# Inter-rater reliability of the Infectious Disease Modeling Reproducibility Checklist (IDMRC) as applied to COVID-19 computational modeling research

**DOI:** 10.1101/2023.03.21.23287529

**Authors:** Darya Pokutnaya, Willem G Van Panhuis, Bruce Childers, Marquis S Hawkins, Alice E Arcury-Quandt, Meghan Matlack, Kharlya Carpio, Harry Hochheiser

## Abstract

**Background:** Infectious disease computational modeling studies have been widely published during the coronavirus disease 2019 (COVID-19) pandemic, yet they have limited reproducibility. Developed through an iterative testing process with multiple reviewers, the Infectious Disease Modeling Reproducibility Checklist (IDMRC) enumerates the minimal elements necessary to support reproducible infectious disease computational modeling publications. The primary objective of this study was to assess the reliability of the IDMRC and to identify which reproducibility elements were unreported in a sample of COVID-19 computational modeling publications.

**Methods:** Four reviewers used the IDMRC to assess 46 preprint and peer reviewed COVID-19 modeling studies published between March 13^th^, 2020, and July 31^st^, 2020. The inter-rater reliability was evaluated by mean percent agreement and Fleiss’ kappa coefficients (κ). Papers were ranked based on the average number of reported reproducibility elements, and average proportion of papers that reported each checklist item were tabulated.

**Results:** Questions related to the computational environment (mean κ = 0.90, range = 0.90–0.90), analytical software (mean κ = 0.74, range = 0.68–0.82), model description (mean κ = 0.71, range = 0.58–0.84), model implementation (mean κ = 0.68, range = 0.39–0.86), and experimental protocol (mean κ = 0.63, range = 0.58–0.69) had moderate or greater (κ > 0.41) inter-rater reliability. Questions related to data had the lowest values (mean κ = 0.37, range = 0.23–0.59). Reviewers ranked similar papers in the upper and lower quartiles based on the proportion of reproducibility elements each paper reported. While over 70% of the publications provided data used in their models, less than 30% provided the model implementation.

**Conclusions:** The IDMRC is the first comprehensive, quality-assessed tool for guiding researchers in reporting reproducible infectious disease computational modeling studies. The inter-rater reliability assessment found that most scores were characterized by moderate or greater agreement. These results suggests that the IDMRC might be used to provide reliable assessments of the potential for reproducibility of published infectious disease modeling publications. Results of this evaluation identified opportunities for improvement to the model implementation and data questions that can further improve the reliability of the checklist.

## Background

Throughout the coronavirus disease 2019 (COVID-19) pandemic, policy makers relied extensively on epidemiological, biostatistical, and computational infectious disease models to inform decisions regarding public health interventions (1). Although there was increased interest in transparent research prior to the pandemic (2), increasingly complex modeling methods and frequently insufficiently detailed descriptions of those methods have led to increasing reproducibility concerns. We recently proposed the Infectious Disease Modeling Reproducibility Checklist (IDMRC) a comprehensive set of guidelines that researchers can follow to publish reproducible modeling results (3). Our goal in this paper is to assess the reliability of the IDMRC to facilitate the reporting of elements impacting the reproducibility of COVID-19 research.

Reproducibility is a cornerstone of the scientific method, enabling the verification of discoveries and protecting against scientific misconduct (4). However, the rapid pace of COVID-19 research has raised concerns about the reproducibility of modeling results. For years governing bodies have published advice to enhance reproducibility of scientific research and proposed lists of elements that should be included in publications to ensure reproducibility have been reported (5–9). Prior to our work, these initiatives have not been synthesized into reliable guidelines for infectious disease computational modeling research. We filled this critical gap in the literature by creating a framework for the implementation of reproducible computational infectious disease models. We formatted the framework into the Infectious Disease Modeling Reproducibility Checklist (IDMRC), a checklist that is applicable to varying types of infectious disease models with ranging complexities (3).

Previously developed guidelines, such as the Strengthening the Reporting of Observational Studies in Epidemiology (STROBE) checklist and the EPIFORGE 2020 guidelines for epidemic forecasting have been instrumental in enhancing the quality of modeling research (10,11). However, they focus on general recommendations for describing elements in a publication without including specific items related to data, analytical software, operating systems (including both names and version numbers), and other key computational components used to conduct the analyses. The IDMRC overcomes these limitations through the inclusion of specific items relevant to publishing reproducible infectious disease modeling studies. Here, we assess the reliability of the IDMRC with multiple reviewers and a sample of COVID-19 computational modeling studies. To our knowledge, this is the first time the reliability of a checklist used to assess the reproducibility of infectious disease modeling studies has been evaluated.

The Models of Infectious Disease Agent Study (MIDAS) Coordination Center (midasnetwork.us) is an NIGMS-funded center supporting the infectious disease research community. Four researchers from the MIDAS Coordination Center evaluated the reliability of the checklist by assessing a random selection of preprint and peer-reviewed COVID-19 modeling papers published between March 13^th^, 2020, and July 31^st^, 2020. The purpose of this study was to assess the inter-rater reliability of the IDMRC, characterize papers based on the reviewers’ qualitative rankings, and determine which reproducibility elements are frequently included or overlooked in COVID-19 computational modeling studies.

## Methods

The IDMRC was previously developed as a framework for the implementation of reproducible computational infectious disease models (Additional File 1) (3). The IDMRC consists of twenty-two questions grouped into six categories: computational environment, analytical software, model description, model implementation, data, and experimental protocol (Additional File 2). We evaluated the performance of the IDMRC in the COVID-19 modeling literature by measuring the agreement among four reviewers for the overall instrument and for individual questions. Based on the evaluations, we made suggested changes to the IDMRC (Additional File 3).

We searched PubMed, medRxiv, arXiv, and bioRxiv using queries for COVID-19 modeling papers between March 13^th^, 2020, and July 31^st^, 2020 (Additional File 4). As preprint servers were widely used to disseminate COVID-19 models at the beginning of the pandemic (12), we included medRxiv, arXiv, and bioRxiv in our search. We dd not restrict to certain types of computational modeling studies in our assessment given that our checklist should be applicable to all computational infectious disease modeling studies ranging from regression models to complex agent-based models. From the search results, we randomly selected 100 papers for title and abstract review (Figure 1).

**Figure 1.** Publications included in the inter-rater reliability analysis of the Infectious Disease Modeling Reproducibility Checklist. *Abbreviations:* COVID-19, coronavirus disease 2019

Four researchers (DP, AAQ, KC, MM) used the IDMRC to independently review the 46 modeling papers to assess which IDMRC elements were included. All four reviewers had experience in reading modeling papers, including training of at least a Master’s in Public Health Degree. DP and AAQ were involved with the development of the checklist and had more experience using the IDMRC relative to KC and MM.

We performed an inter-rater reliability analysis to assess the concordance of the ratings of the ordinal categorical items. Reliability was assessed based on the mean percent agreement with Wald 95% confidence intervals and Fleiss’ kappa (κ) estimates. Fleiss’ kappa is the observed agreement corrected for the agreement expected by chance and is appropriate when there are more than two raters assessing ordinal or nominal data (13). We used the Power4Cats function in the kappaSize package in R version 4.0.2, RStudio Version 1.3.107 to determine that 46 publications could reliably produce a lower limit for a kappa estimate of 0.293 (1). Fleiss’ kappa was computed with linear weights using the wlin.conc function in the R raters package 2.0.1 (14,15). Monte Carlo simulations were used to calculate percentile bootstrap confidence intervals. Results were interpreted using previously published guidelines: κ < 0.01 indicates no agreement; κ = 0.01–0.20, slight; κ = 0.21–0.40, fair; κ = 0.41–0.60, moderate; κ = 0.61–0.80 substantial; and κ = 0.81–1 almost perfect agreement (15). A kappa score below 0.41 falls into the category of “slight” agreement and was deemed by the authors as indicative of questions that needed to be reviewed and revised.

For each reviewer, we qualitatively ranked the papers by the number of reported elements in each publication. Publications with the most elements included (as rated by the reviewers) were ranked the highest. We also averaged the reviewers’ rankings to report the average qualitative rankings of the 46 publications. To assess if the potential impact of the peer-review on the number of reproducibility elements, DP independently reviewed the five highest-rated and 5 lowest-rated publications to determine 1) if any of the publications that were published in preprint servers at the time of the review had since been published in peer-reviewed journals, and 2) if the papers that had been published in peer-reviewed journals had reported more reproducibility elements in those publications. Finally, we tabulated the proportion of papers that reported each checklist element as well as the proportion of checklist elements reported in all publications (both averaged across all reviewers).

## Results

Four MIDAS researchers used the IDMRC to review 46 COVID-19 computational modeling papers published between March 13^th^, 2020, and July 31^st^, 2020. After title and abstract review, 48 papers were excluded based on the following exclusion criteria: observational, genomic, immunological, and molecular studies, commentaries, reviews, retractions, letters to editor, response papers, papers not related to COVID-19, and descriptions of software applications. Of the remaining 48 papers, two publications reviewing previously developed COVID-19 models were excluded after full text review (Figure 1). The final 46 paper sample consisted of 39 (85%) publications published in preprint servers (n = 34 from medRxiv; n = 5 from arXiv) and 7 (15%) from peer-reviewed journals (Additional File 5).

### Inter-rater reliability of the IDMRC

The inter-rater reliability evaluation indicated that the IDMRC was a reliable tool with most questions characterized by moderate or better (κ > 0.41) agreement between the four reviewers. Overall, the mean percent agreement ranged from 54% (data question 5.3) to 94% (computational environment 1.1, 1.2; model implementation 4.6). Fleiss’ kappa estimates ranged from 0.23 (95%CI 0.10, 0.40) for IDMRC data question 5.5 to 0.90 (95%CI 0.79, 0.98) for both computational environment questions. Several Fleiss’ kappa estimates in the model implementation and data categories fell below moderate agreement (Table 1).

**Table 1.**
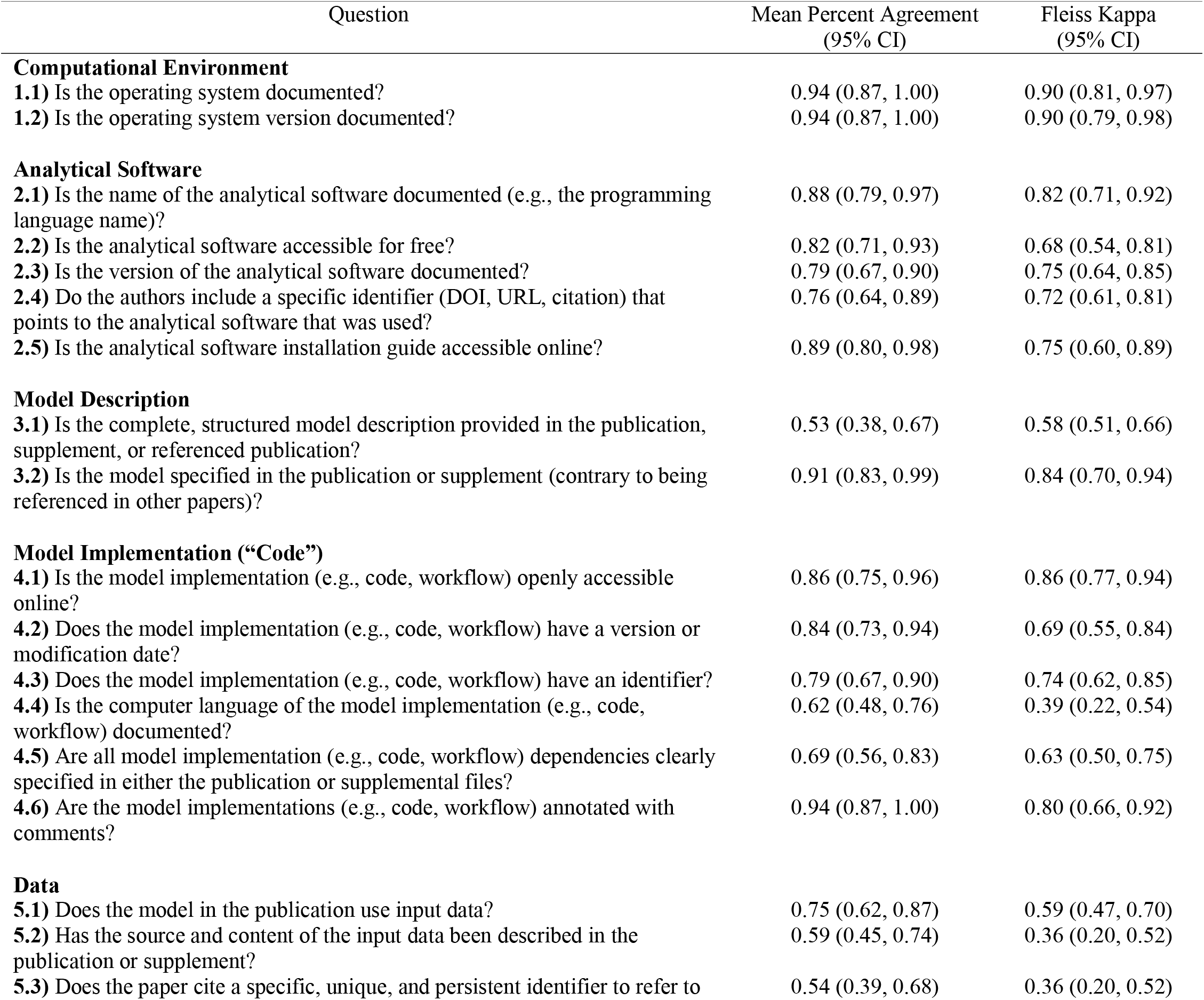
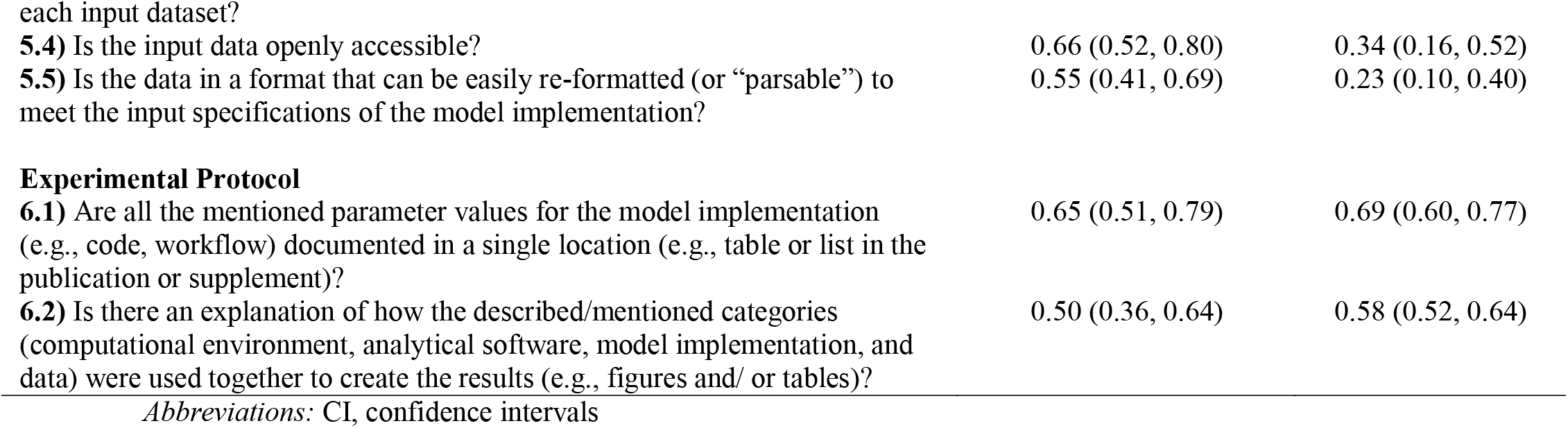
Infectious Disease Modeling Reproducibility Checklist elements reported in COVID-19 modeling papers.

### Characterization of papers based on reviewer qualitative rankings

Reviewers identified similar publications as reporting the most reproducibility elements (i.e., reviewers reported “yes” for more questions) or the least number of elements (i.e., reviewers reported “yes” less often). KC and MM, the two reviewers with the least experience using the IDMRC, agreed upon eight publications in the top 25% (n = 13) and the eight publications in the bottom 25% (n = 13) (Additional File 6). DP and AAQ, the two reviewers with more experience using the checklist, agreed on nine publications in the top 25% and ten publications in the bottom 25% (Additional File 6). All four reviewers agreed on six publications in the top 25% and seven publications were reported in the bottom 25% (Additional File 7). The publications with the most reproducibility elements based on average scores (publications 9, 15, 16, 19, and 27) and the least reported reproducibility elements (2, 12, 20, 23, and 37) were all originally published as preprints. Four publications (2, 19, 20, and 27) have since been published in peer-reviewed journals. An independent review of these four papers by DP determined that the peer-reviewed versions did not have a significantly increased number of reported reproducibility elements.

### Average proportion of papers that reported each checklist item

Rates of inclusion of the 22 checklist elements varied from 2% of papers reporting the operating system version (question 1.2) to 92% providing the model description in the journal or publication as opposed to referencing a previously developed model (question 3.2, Figure 3A). Fifty percent (n = 23) of publications provided less than 40% of all checklist categories (Figure 3B). Over 94% of studies did not provide either the name or the version of the operating system used in their analysis (questions 1.2, 1.3, respectively) (Figure 3A). The analytical software name (e.g., R, STATA, SAS) was provided in 62% of publications (question 2.1), but only 41% of the software tools were openly accessible without a licensing fee (question 2.2). Most studies provided the input data (70%; question 5.4); however, only 25% provided the model implementation, or code, used to generate the data (question 4.1). Averaged across the raters, over 50% of the publications provided all five data elements (questions 5.1−5.5), but less than 50% of the publications provided all six model implementation elements (questions 4.1−4.6). Thirty-nine percent of publications provided the parameters used in their models (questions 6.1) while 19% provided a clear explanation for how categories 1−5 were used together to create the model results (question 6.2).

**Figure 2.** Average quantitative paper ranking (n = 46) among four reviewers. Green bars correspond to the average number of reported elements in each publication (“yes” responses); yellow indicates partially reported elements; red indicates not reported elements, gray indicates not applicable responses.

**Figure 3.** Proportion of coronavirus disease 2019 (COVID-19) modeling publications (n = 46) that reported each infectious disease modeling reproducibility checklist (IDMRC) component elements. A, average percentage of papers that reported each checklist element; B, average proportion of checklist elements that were reported in all publications. Dashed line in B indicates the mean.

## Discussion

Improved reproducibility of infectious disease computational models will help researchers efficiently build upon previous studies and accelerate the pace of scientific advancements. We previously developed the Infectious Disease Modeling Reproducibility Checklist (IDMRC) to enumerate the elements necessary to support reproducible infectious disease computational modeling studies (3). Our evaluation indicated that the IDMRC is a reliable tool with the majority of the inter-rater reliability estimates reporting moderate or greater agreement between the four reviewers. Participating reviewers placed similar publications in the top and bottom reproducibility score quantiles based on the number of elements missing in each publication. The two experienced and the two novice checklist users had more similar rankings, suggesting that formally training researchers to use the IDMRC prior to evaluating a study may produce more consistent results. Furthermore, revisions to the checklist questions, primarily in the model implementation and data sections, may increase reliability of future evaluations.

Our experience with the application of the checklist suggests that the question ordering may have impacted reliability. The question regarding whether the model implementation computer language was documented (question 4.4) may have received a lower score due to its positioning after the analytical software name question (question 2.1). In most instances if a publication reported the analytical software name (e.g., R, STATA), the model implementation computer language would be evident (e.g., R uses R coding language, STATA uses STATA coding language). However, occasionally the two may differ, such as when researchers use their own developed software or utilize packages to develop scripts in languages that are not original to the analytical software (e.g., writing Python scripts in R with the use of the *reticulate* package). Additionally, if a reviewer had already selected “no” or “not applicable” for prior a model implementation question (questions 4.1−4.3), the reviewer may have automatically selected the same response for question 4.4 without independent thought to the question. Moving the question from the model implementation section to the analytical software section (after question 2.1) could improve the checklist reliability. We propose a revised version of the checklist which includes question 4.4 directly after question 2.1 (Additional File 3).

Reliability assessments can also help highlight ambiguities in definitions commonly used in infectious disease computational modeling literature. Lower κ estimates in the data section may have been due to uncertainty regarding the definition of input data (question 5.1). For example, some of the publications described susceptible-infected-recovered (SIR) compartmental models which can be parametrized using input data, simulated data, or by referencing previously reported parameters. In these situations, reviewers may have not considered parameters as input data. To improve the reliability of the checklist, we defined input data as “any data, including parameters, used to generate a model or initial conditions” in the updated version of the checklist (Additional File 3). Furthermore, we originally included a “not applicable” answer choice in question 5.1; however, after the reliability assessment, we deemed that this answer choice was not warranted because a response of “yes” or “no” should capture all possible answer choices. Thus, we removed the “not applicable” answer choice from question 5.1 in the newest version of the checklist (Additional File 3). Given that conditional nature of the checklist questions (i.e., subsequent checklist questions are affected by prior responses), including a definition of the input data as well as correcting the answer choices in question 5.1 could improve the reliability of the following data questions.

The computational environment, which comprises the operating system name and version, was least reported. Failure to reproduce modeling studies, even if the data and code have been made available, can be due to incompatibilities or specific requirements in the computational environment (16). For example, SAS software is not compatible with the macOS operating system unless it is run in a virtual machine. Software developed by the authors of a given paper or analyses that require high-performance computing may also require specific types of operating systems to be functional. We recommend including a short statement specifying the name and version of the operating system in future infectious disease computational modeling studies.

Two of the top qualitatively ranked publications (19 and 27) as well as two of the lower ranked publications (2 and 20) were initially published in medRxiv, during the time of review, but have since been published in peer reviewed journals. An independent review by DP indicated that the peer reviewed versions of these paper did not include significant improvement in the number of reported reproducibility elements. This suggests that the peer review process does not necessarily improve the reproducibility of papers in our sample. Despite an increase in the adoption of data and code sharing policies by journals, stricter application of the IDMRC or similar guidelines may be needed to further improve reproducibility during the peer review process (17). Some suggestions include the complementary submission of checklists, such as the IDMRC, or dynamic computational notebooks (17,18).

Over 70% of the publications provided the data used for their analysis. Our estimate was similar to the 60% (n = 29) of CDC-compiled COVID-19 modeling studies analyzed by Jalali et al. and much higher than the 24.8% (n = 332) reviewed by Ioannidis et al. that reported to share their data. However, Ioannidis et al., used a text mining algorithm which may not have picked up publications that shared their data (19,20). Many journals now require researchers to provide a data availability statement when submitting a publication but allow researchers to circumnavigate the provision by stating “the datasets and code are available from the corresponding author on reasonable request.” Some publishers require authors to make their publication data publicly available (21). As of January 25, 2023, National Institute of Health-supported research requires researchers to include a plan for data sharing within their funding applications. While our review included publications published on preprint servers, which have less strict reporting guidelines, we reason that preprint COVID-19 computational models should have been just as transparent with their data as peer-reviewed publications given their widespread use by policymakers and news outlets during the start of the pandemic (12).

Although providing data access is becoming a common practice in infectious disease computational studies, progress in sharing model implementations is lagging. In our sample, most papers provided the model description; however, the code used to implement the model and create the results was reported in less than 25% of the studies. In the previous review of COVID-19 computational modeling studies, researchers found that a similar 21.5% of publications reported the code (n = 288) (20). With increasingly complex computational methodologies in infectious disease modeling literature, withholding the exact data manipulation and analysis steps can impede the consistent regeneration of modeling results. Researchers should aim to provide open-source access to appropriately versioned model implementations accompanied by comprehensible annotations in online repositories.

Sharing a reproducible model consists of more than just sharing the data or code. Each component in the checklist works together to produce the final modeling result. With each additional missing component, the time and effort that it takes for future reproduction attempts increases (22). Amid a pandemic, timely, reproducible research is critical in informing policies and life-saving interventions.

The present study has several limitations. First, we sampled COVID-19 computational modeling studies published early in the pandemic when authors may have reported fewer reproducibility elements compared to publications published in later periods. In future work, assessing the reproducibility of publications reported during various stages of the pandemic may lead to insights regarding timing of publications and reproducibility of modeling literature.

Second, given that two of the reviewers had limited experience using the IDMRC prior to the assessment, we may have underestimated the true reliability of the IDMRC. Furthermore, differences in reviewer experience may have led to an under- or over-estimation of the average number of reported reproducibility elements in our sample of publications. Finally, while the reliability assessment of the IDMRC goes a step beyond most checklists, we did not assess the reliability of the proposed changes to the IDMRC.

## Conclusions

Our review focused on evaluating the performance of the IDMRC in COVID-19 computational modeling publications. Additional rounds of review with more reviewers and modeling studies outside of COVID-19 might generalize reliability. Furthermore, lower inter-rater reliability scores on some of the elements may have impacted the reported frequencies of missing reproducibility elements. To address these issues, we proposed a revised version of the IDMRC. Tools such as the IDMRC can encourage the documentation and sharing of all elements necessary to reproduce a computational modeling study, thus supporting reproducible computational infectious disease studies and accelerating scientific discoveries by allowing others to validate results as well as by providing resources that might be reused in future studies.

## Data Availability

All data produced in the present work are contained in the manuscript.

## Declarations

### Availability of data and materials

All data generated or analyzed during this study are included in this published article and its supplementary information files.

### Authors’ contributions

Conceptualization: DP, BC, WVP; Methodology: DP, BC, WVP; Software: DP; Validation: DP; Formal analysis: DP; Investigation: DP, BC, WVP, AAQ, MM, KC; Data curation: DP; Writing – original draft: DP, BC, WVP; Writing – review & editing: DP, BC, WVP, HH, MH, AAQ, MM, KC, MR; Visualization: DP, BC, WVP, HH; Supervision: BC, WVP, HH; Project administration: WVP, HH; Funding acquisition: WVP

## Acknowledgements

We thank current and past members of the Public Health Dynamics Lab, including and Dr. Mark S. Roberts, Jessica Kerr, Lucie Contamin, Anne Cross, John Levander, Jeffrey Stazer, Inngide Osirus, and Lizz Piccoli for critical discussions and feedback. We would also like to thank Dr. Anne Newman for her feedback during the early development stages of this manuscript.

