## Additional File 1 for "Inter-rater reliability of the Infectious Disease Modeling Reproducibility Checklist (IDMRC) as applied to COVID-19 computational modeling research"

**Table S1.** Framework categories and elements and relevant examples.

| Category and Element | Abbreviated Definition* | Examples |
| --- | --- | --- |
| **1) Computational Environment** | | |
| 1.1) Name | Name of operating system | “Microsoft Windows,” “macOS,” “Linux” |
| 1.2) Version | Version of operating system | “Windows 8.1 (Blue),” “macOS 10.12 (Sierra)” |
| **2) Analytical Software** | | |
| 2.1) Name | Name of software program | “SAS,” “R,” “original software name” |
| 2.2) License | Restriction on use | Open source: “R”; propriety: “STATA,” “SAS” |
| 2.3) Version | Version of software | “SAS 9.4,” “R version 3.3.3” |
| 2.4) Identifier | Unique online identifier | “DOI,” “URL” |
| 2.5) Documentation | Availability of documentation to use and install software | URL to installation guide |
| **3) Model Description** |  |  |
| 3.1) Description | Complete, structured description of the model | Equations, diagrams, charts, tables vs. unstructured text |
| 3.2) Location | Specified in the publication/ supplement | Model described in the methods/supplement vs. referenced in preceding publication |
| **4) Model Implementation (“Code”)** | | |
| 4.1) License | Restriction on use | Publicly stored on GitHub |
| 4.2) Version | Version of code | Version, modification date |
| 4.3) Identifier | Unique online identifier | “DOI,” “URL” |
| 4.4) Computer language | Name of code language | “SAS,” “R,” “STATA,” “Python,” “C++” |
| 4.5) Dependencies | Additional essential files | Packages, classes, supplementary files |
| 4.6) Annotations | Sufficient, user-interpretable comments | Code suitably annotated for understanding |
| **5) Data** | | |
| 5.1) Data calibration | Indicator for whether the model was calibrated to existing vs. simulated data | “Yes” (model calibrated to input data); “no” (model does not require input data) |
| 5.2) Definition | Description of data content and source | Content: column/ field descriptions; source: “CDC,” “Project Tycho” |
| 5.3) Identifier | Identifier from where the data are retrieved | “DOI,” “URL” |
| 5.4) License | Restriction on use | Publicly stored on GitHub |
| 5.5) Data format | Formatted data for the model implementation | “CSV,” “JSON,” “XML” |
| **6) Experimental Protocol** | | |
| 6.1) Parameters | Description of model parameters | Table or list of parameter values |
| 6.2) Scientific workflow | Process of how categories 1-5 create the results | Explanation in GitHub README.md |
