## Additional File 2 for "Inter-rater reliability of the Infectious Disease Modeling Reproducibility Checklist (IDMRC) as applied to COVID-19 computational modeling research"

**Table S2. Original Infectious Disease Modeling Reproducibility Checklist with examples.** The checklist consists of the computational environment, analytical software, model description, model implementation, data, and experimental protocol and their associated elements. Each question corresponds to several answer options. Additional examples for each element are provided.

| **1. Computational Environment** | | |
| --- | --- | --- |
| 1.1) Is the operating system documented? | Examples: Microsoft Windows, macOS, Linux | ▢ Yes; the operating system is  documented  ▢ No; the operating system is not documented  ▢ Not applicable |
| 1.2) Is the operating system version documented? | Examples: Windows 8.1 (Blue) or macOS 10.12 (Sierra) | ▢ Yes; the operating system version is documented  ▢ No; the operating system version is not documented  ▢ Not applicable |
| **2. Analytical Software** | | |
| 2.1) Is the name of the analytical software documented (e.g., the programming language name)? | Examples: SAS, R, STATA, Python, C++. Authors may have also used an originally developed software with a unique name. | ▢ Yes; the name of the analytical software is documented  ▢ No; the name of the analytical software is not documented  ▢ Not applicable |
| 2.2) Is the analytical software accessible for free? | For all mentioned analytical software, if the analytical software is available online for download and it is free, mark "Yes." If the analytical software is available online but must be bought or licensed, select "Partially." Additionally, if the authors used multiple types of analytical software in their analyses and not all of them are available for free download, select "Partially." If the analytical software is not available online for download, select "No." Examples of analytical software available for free download: Java, R, Python; non-free examples: STATA, SAS, SPSS, MATLAB. | ▢ Yes; all mentioned software is available for free download  ▢ Partially; all mentioned software is available for download but requires a purchase and/or acquiring a license or not all the software used were available for free download  ▢ No; the software is not available for download  ▢ Not applicable |
| 2.3) Is the version of the analytical software documented? | For all mentioned analytical software, if the version is documented in the publication, supplement, or in an online repository (e.g., GitHub), select "Yes." If the version is documented for some of the mentioned analytical software, select "Partially." If the version is not documented, select "No." Examples: SAS 9.4 or R version 3.3.3. | ▢ Yes; the version is documented for all mentioned analytical software  ▢ Partially; the version is documented for some mentioned analytical software  ▢ No; the version of the analytical software is not documented  ▢ Not applicable |
| 2.4) Do the authors include a specific identifier (DOI, URL, citation) that points to the analytical software that was used? | If the authors provide a reference for all mentioned software that includes a DOI, URL, or version from which the software was retrieved from, select "Yes." If all mentioned software is referenced but the reference does not include the DOI, URL or version, select "Partially." Additionally, if only some mentioned software is referenced, select “Partially.” If the authors do not provide a reference for the software, select “No.” | ▢ Yes; all mentioned software is referenced with a specific identifier such as a DOI, URL, or version  ▢ Partially; all mentioned software is referenced but the reference does not include the DOI, URL, or version or only some mentioned software is referenced  ▢ No; none of the mentioned software is referenced  ▢ Not applicable |
| 2.5) Is the analytical software installation guide accessible online? | For all mentioned analytical software, if the publication includes a link to the installation guide or the installation guide is accessible online, select "Yes." If the installation guide is not accessible online, select "No." Commonly used analytical software such as Java, R, Python, STATA, SAS, SPSS, MATLAB, SAS, STATA have accessible online installation guides. If the authors used an originally developed analytical software, the software may or may not have an accessible installation guide. | ▢ Yes; for all mentioned analytical software, the installation guides are accessible online  ▢ Partially; for some of the mentioned analytical software, the installation guides are accessible online  ▢ No; for all mentioned analytical software, the installation guides are not accessible online  ▢ Not applicable |
| **3. Model Description** | | |
| 3.1) Is the complete, structured model description provided in the publication, supplement, or referenced publication? | If there is a complete, structured model description in (e.g., figure, table, or text) in the publication, supplement, or in a referenced publication in a single location, select "Yes." If there are portions of the structured model description throughout the publication, supplement, or referenced publication, but the portions together seem complete, select "Partially." If there is an unstructured but complete model description in the publication, supplement, or referenced publication, select “Partially.” If there is an incomplete or no model description, select "No." | ▢ Yes; there is a complete, structured model description in the publication, supplement, or referenced publication  ▢ Partially; there are portions of the structured model description throughout the publication, supplement, or referenced publication, but the portions together seem complete  ▢ Partially; there is an unstructured but complete model description  ▢ No; there is an incomplete or no model description  ▢ Not applicable |
| 3.2) Is the model specified in the publication or supplement (contrary to being referenced in other papers)? | If the authors used a structured or unstructured model that they described in the publication or its supplemental materials, select "Yes." If the authors cited a previously developed model without describing the model in their publication, select "No." | ▢ Yes; there is a model specified in the publication or supplement  ▢ No; the authors cited a previously developed model in a referenced paper  ▢ Not applicable |
| **4. Model Implementation (“Code”)** | | |
| 4.1) Is the model implementation (e.g., code, workflow) openly accessible online? | If the model implementation (e.g., code, workflow) is provided such that all results (e.g., figures, tables) in the publication can be recreated, select "Yes." A workflow can include instructions of how to configure/use a graphical user interface to recreate published results. If the implementation is partially provided, such that only part of the results can be recreated select "Partially." If the model implementation is not provided, select "No." | ▢ Yes; the model implementation for all results is openly accessible online  ▢ Partially; the model implementation for part of the results is openly accessible online  ▢ No; the model implementation is not openly accessible online  ▢ Not applicable |
| 4.2) Does the model implementation (e.g., code, workflow) have a version or modification date? | If the version or modification date for the model implementation is documented in the publication, supplement, or in an online repository (e.g., GitHub), select "Yes." If the version or modification date is not provided or it is unclear as to which model implementation the author's used, select, "No." | ▢ Yes; the version or modification date is documented in the publication, supplement, or in an online repository (e.g., GitHub)  ▢ No; the version or modification date is not provided, or it is unclear as to which model implementation the author's used  ▢ Not applicable |
| 4.3) Does the model implementation (e.g., code, workflow) have an identifier? | If the model implementation has a unique, persistent, and specific identifier (e.g., DOI or URL to a specific file) or the model implementation is provided in the paper or supplement, select "Yes." If there is a general URL to the model implementation (e.g., a URL to an entire GitHub repository), select "Partially." If the model implementation has no identifier, select, "No." If the model implementation is not available, then select "Not applicable." | ▢ Yes; there is a unique, persistent, and specific identifier (e.g., DOI or specific URL) for the model implementation or the model implementation is provided in the paper or supplement  ▢ Partially; there is a general URL for the model implementation  ▢ No; there is no identifier for the model implementation  ▢ Not applicable |
| 4.4) Is the computer language of the model implementation (e.g., code, workflow) documented? | Often the language of the model implementation will be unambiguous. For example, R uses model implementations written in R language and SAS uses model implementations written in SAS language; however, some publications may use originally developed analytical software. In these situations, it is important for the authors to specify what language(s) the program uses. If the computer language for the model implementation is documented, select "Yes." If the computer language for the model implementation is not documented, select "No." | ▢ Yes; the computer language of the model implementation is documented  ▢ No; the computer language of the model implementation is not documented  ▢ Not applicable |
| 4.5) Are all model implementation (e.g., code, workflow) dependencies clearly specified in either the publication or supplemental files? | In some cases, the model implementation may have additional dependencies (e.g., packages or classes). For example, the author's may have imported dplyr or ggplot2 packages in R or the Scanner class in Java in order run the model. Even if the authors do not provide the model implementation, they may still mention the dependencies they used in the methods section. Analytical software that commonly use packages that are not pre-installed include Java, R, and Python. Analyses in SAS, MATLAB, and STATA occasionally require additional model implementation dependencies. SPSS does not have additional model implementation dependencies. If all the model implementation dependencies are clearly specified in either the publication, supplemental files, or at the top of the model implementation, select "Yes." If some of the model implementation dependencies are listed in the publication, supplemental files, or are dispersed throughout the model implementation, select "Partially." If none of the model implementation dependencies are specified, select "No." | ▢ Yes; all the model implementation dependencies are clearly specified in either the publication, supplemental files, or at the top of the model implementation  ▢ Partially; some of the model implementation dependencies are listed in the publication, supplemental files, or are dispersed throughout the model implementation  ▢ No; none of the model implementation dependencies are specified  ▢ Not applicable |
| 4.6) Are the model implementations (e.g., code, workflow) annotated with comments? | If the model implementation is annotated with comprehensible comments that allow the user to easily interpret what the code is doing, select "Yes." If there are no comments accompanying the model implementation, select "No." | ▢ Yes; the model implementations are annotated with comprehensible comments  ▢ No; the model implementations are not annotated with comments  ▢ Not applicable |
| **5. Data** | | |
| 5.1) Does the model in the publication use input data? | Some models, such as stochastic simulation models, may not require input data. If you answer, "No", you will skip to the next section about the experimental protocol. | ▢ Yes; the model uses input data  ▢ No; the model does not require input data  ▢ Not applicable |
| 5.2) Has the content and source of the input data been described in the publication or supplement? | If any of the content (e.g., variable descriptions, number of observations) and source information (e.g., data was from Johns Hopkins, CDC, specific websites, interviews) were described in the publication or supplement for each input dataset, select "Yes." If any of the content and source information was described in the publication or supplement for some input datasets, select "Partially." If any of the content or source information was described for each input dataset, select “Partially.” If neither source nor content was described for each input dataset, or only the source or content described for some input datasets, select “No.” | ▢ Yes; the source and content have been described for each input dataset  ▢ Partially; the source and content have been described for some input datasets  ▢ Partially; the source or content have been described for each input dataset  ▢ No; neither the source nor content was described for each input dataset, or only the source or content described for some input datasets  ▢ Not applicable |
| 5.3) Does the paper cite a specific, unique, and persistent identifier to refer to each input dataset? | If each input dataset has a unique, persistent, and specific identifier (e.g., a DOI or a URL to a specific file or set of files), select "Yes." A specific URL would point to a data file, a version, and/or specific download date to enable re-downloading of the same data used in the analysis. Additionally, if the data are provided in the publication or supplement itself, select "Yes." If there is a general URL to each input dataset (e.g., a URL to a GitHub repository but that it is unclear as to which files are used in the analysis or a web-based data archive), select "Partially." If only some of the input datasets have a unique, persistent, and specific identifier or a general URL, select "Partially." If none of the data described have an identifier, select, "No." | ▢ Yes; there is a unique, persistent, and specific identifier (e.g., DOI or specific URL to a specific file or set of files), or  the data are included in the paper or supplement  ▢ Partially; there is a general URL (e.g., a URL to a GitHub repository or online data archive) for data or used in the analysis  ▢ Partially; some of the input data have unique, persistent, and specific identifiers  ▢ No; none of the input data have an identifier  ▢ Not applicable |
| 5.4) Is the input data openly accessible? | If all the mentioned data used to create the results (e.g., figures and/ or tables) are openly accessible in either the publication, supplement, a specified online repository (e.g., GitHub), or can be downloaded from a specified website (e.g., Johns Hopkins dashboard, CORD-19, Project Tycho), select “Yes.” If some of the mentioned data used to create the results are provided in either the publication, supplement, online repository, or can be downloaded from a website, select "Partially." If the data are not openly accessible, or if the authors state that the data are not available, select, "No." | ▢ Yes; all the mentioned data used in the analysis are openly accessible in either the publication, supplement, online repository, or can be downloaded from a website  ▢ Partially; part of the mentioned data used in the analysis are openly accessible in either the publication, supplement, online repository, or can be downloaded from a website  ▢ No; none of the mentioned data used to create the results are openly accessible  ▢ Not applicable |
| 5.5) Is the data in a format that can be easily re-formatted (or “parsable”) to meet the input specifications of the model implementation? | If all the mentioned input data are in a format that can be easily re-formatted (e.g., CSV, JSON, XML, PDF table that can be copied as a CSV), select "Yes." If some of the mentioned input data are in a format that can be easily re-formatted, select "Partially." If the data are in a format that is not easily re-formatted (e.g., narrative text, PDF image file, PNG), select "No." | ▢ Yes; all the input data are in a format that can be easily re-formatted  ▢ Partially; some of the input data are in a format that can be easily re-formatted  ▢ No; all data are in a format that is not easily re-formatted  ▢ Not applicable |
| **6. Experimental Protocol** | | |
| 6.1) Are all the mentioned parameter values for the model implementation (e.g., code, workflow) documented in a single location (e.g., table or list in the publication or supplement)? | If the authors provide the parameter values for the model implementation in a single location (e.g., table or list) in the text or supplement, select “Yes”. If the authors provide some of the parameter values or the parameter values are dispersed throughout the text or supplement, select “Partially”. It the authors do not provide any of the parameter values, select “No”. | ▢ Yes; all mentioned parameter values for the model implementation are documented in a single location  ▢ Partially; some mentioned parameters for the model implementation are documented or they are dispersed throughout the text  ▢ No; none of the parameter values for the model implementation are documented  ▢ Not applicable |
| 6.2) Is there an explanation of how the described/mentioned categories (computational environment, analytical software, model implementation, and data) were used together to create the results (e.g., figures and/ or tables)? | If there is a complete explanation in the publication, supplement, or online repository for how all the categories (computational environment, analytical software, model description, model implementation, data) were used to create the results, select “Yes.” If there is a partial explanation in the publication, supplement, or online repository for how all the mentioned were used to create the results, select “Partially.” If there is no explanation of how the mentioned categories were used together to create the results, select "No." If the authors did not mention more than one of the previous categories, select "Not applicable. | ▢ Yes; there is complete explanation in the publication, supplement, or online repository for how all the mentioned categories were used to create the results  ▢ Partially; there is a partial explanation or how the data and model implementation were used to create the key results  ▢ No; there is not an explanation of how the data and model implementation were used to create the key results  ▢ Not applicable |
