## Additional File 4 for "Inter-rater reliability of the Infectious Disease Modeling Reproducibility Checklist (IDMRC) as applied to COVID-19 computational modeling research"

**Table S4.** Search terms used to query PubMed, medRxiv, bioRxiv, and arXiv for the subset of COVID-19 modeling papers published between March 13^th^, 2020, and July 31^st^, 2020.

| PubMed | medRxiv, bioRxiv, arXiv |
| --- | --- |
| (\"coronavirus\" [MeSH Terms] OR coronavirus[Title/Abstract] OR COVID-19[Title/Abstract] OR 2019-nCoV[Title/Abstract] OR SARS-CoV-2[Title/Abstract]) AND (estimat*[Title/Abstract] OR model*[Title/Abstract] OR reproduct*[Title/Abstract]) NOT (review[Title] NOT systematic[Title] NOT meta-analysis[Title] NOT patients[Title]) | (ti:covid OR ti:coronavirus OR ti:2019-ncov OR ti:SARS-CoV-2 OR abs:covid OR abs:coronavirus OR abs:2019-ncov OR abs:SARS-CoV-2) AND (ti:model* OR abs:model* OR ti:estimat* OR abs:estimat* OR ti:reproduct* OR abs:reproduct*) AND lastUpdatedDate:["+start_date_str + " TO " + end_date_str+"] ANDNOT(ti:review OR ti:systematic OR ti:meta-analysis OR ti:patients) |
