## Additional File 5 for "Inter-rater reliability of the Infectious Disease Modeling Reproducibility Checklist (IDMRC) as applied to COVID-19 computational modeling research"

**Table S5.** Paper identifier, original DOI and source at the time of review, and average number of items reported (i.e., reviewers reported “yes”) for the forty-six-coronavirus disease 2019 computational modeling papers assessed using the Infectious Disease Modeling Reproducibility Checklist. For papers published in pre-print servers at the time of review, the DOI of the subsequent peer-reviewed paper is included, in applicable.

| Paper  Identifier | Publication title | Original DOI | Original Source | Average number of items reported (SD) | Subsequent DOI, if applicable |
| --- | --- | --- | --- | --- | --- |
| 1 | A first study on the impact of containment measure on COVID-19 spread in Morocco | 10.1101/2020.04.26.20080770 | medRxiv | 6.5 (1.91) | https://doi.org/10.1016/j.chaos.2020.110231 |
| 2 | A model showing the relative risk of viral aerosol infection from breathing and the benefit of wearing masks in different settings with implications for Covid-19 . | 10.1101/2020.04.28.20082990 | medRxiv | 1.75 (0.96) | 10.37871/ajeph.id32 |
| 3 | A novel cohort analysis approach to determining the case fatality rate of COVID-19 and other infectious diseases | 10.1101/2020.04.02.20051482 | medRxiv | 5.25 (0.96) | 10.1371/journal.pone.0233146 |
| 4 | A pandemic at the Tunisian scale. Mathematical modelling of reported and unreported COVID-19 infected cases | 10.1101/2020.05.21.20108621 | medRxiv | 7 (2.16) |  |
| 5 | A simulation of a COVID-19 epidemic based on a deterministic SEIR model | 10.1101/2020.04.20.20072272 | medRxiv | 9.25 (0.5) | 10.3389/fpubh.2020.00230 |
| 6 | ARIMA forecasting of COVID-19 incidence in Italy, Russia, and the USA | 10.48550/arXiv.2006.01754 | arXiv | 9.75 (1.5) |  |
| 7 | Asymptotic estimates of SARS-CoV-2 infection counts and their sensitivity to stochastic perturbation | 10.1063/5.0008834 | *Chaos: An Interdisciplinary Journal of Nonlinear Science* | 7.5 (1.91) |  |
| 8 | Bayesian Modeling of COVID-19 Positivity Rate -- the Indiana experience | 10.48550/arXiv.2007.06541 | arXiv | 11 (1.41) |  |
| 9 | Cooperative virus propagation in COVID-19 transmission | 10.1101/2020.05.05.20092361 | medRxiv | 14 (2.83) |  |
| 10 | COVID 19: Real-time Forecasts of Confirmed Cases, Active Cases, and Health Infrastructure Requirements for India and its Majorly Affected States using the ARIMA model. | 10.1101/2020.05.17.20104588 | medRxiv | 9.5 (1.29) |  |
| 11 | COVID-19 in India: State-wise Analysis and Prediction | 10.1101/2020.04.24.20077792 | medRxiv | 4.25 (0.96) | 10.2196/20341 |
| 12 | Dynamics of Interacting Hotspots -- I | 10.48550/arXiv.2004.12799 | arXiv | 2.75 (2.06) |  |
| 13 | Transmission dynamics and control of COVID-19 in Chile, March-June, 2020 | 10.1101/2020.05.15.20103069 | medRxiv | 8.5 (4.04) | 10.1371/journal.pntd.0009070 |
| 14 | Effects of age-targeted sequestration for COVID-19 | 10.1080/17513758.2020.1795285 | *Journal of Biological Dynamics* | 11.5 (3.32) |  |
| 15 | Effects of weather and policy intervention on COVID-19 infection in Ghana | 10.48550/arXiv.2005.00106 | arXiv | 13.75 (1.26) |  |
| 16 | Estimating effects of physical distancing on the COVID-19 pandemic using an urban mobility index | 10.1101/2020.04.05.20054288 | medRxiv | 16.5 (1.73) |  |
| 17 | Estimating the number of SARS-CoV-2 infections and the impact of social distancing in the United States | 10.48550/arXiv.2004.02605 | arXiv | 12 (3.74) |  |
| 18 | Estimating the number of SARS-CoV-2 infections in the United States | 10.1101/2020.04.13.20064519 | medRxiv | 12.25 (2.63) |  |
| 19 | Estimating unobserved SARS-CoV-2 infections in the United States | 10.1101/2020.03.15.20036582 | medRxiv | 13.25 (6.55) | 10.1073/pnas.2005476117 |
| 20 | Evaluation of the secondary transmission pattern and epidemic prediction of COVID-19 in the four metropolitan areas of China | 10.1101/2020.03.06.20032177 | medRxiv | 3.25 (1.26) | 10.3389/fmed.2020.00171 |
| 21 | Extensions of the SEIR Model for the Analysis of Tailored Social Distancing and Tracing Approaches to Cope with COVID-19 | 10.1101/2020.04.24.20078113 | medRxiv | 10.75 (3.3) | 10.1038/s41598-021-83540-2 |
| 22 | Healthcare impact of COVID-19 epidemic in India: A stochastic mathematical model | 10.1016/j.mjafi.2020.03.022 | *Medical Journal Armed Forces India* | 8.5 (4.51) |  |
| 23 | Herd immunity vs suppressed equilibrium in COVID-19 pandemic: different goals require different models for tracking | 10.1101/2020.03.28.20046177 | medRxiv | 3.75 (2.99) |  |
| 24 | Impact Assessment of Full and Partial Stay-at-Home Orders, Face Mask Usage, and Contact Tracing: An Agent-Based Simulation Study of COVID-19 for an Urban Region | 10.1101/2020.07.27.20163121 | medRxiv | 9.25 (4.72) | 10.1016/j.gloepi.2020.100036 |
| 25 | Intervention Serology and Interaction Substitution: Modeling the Role of ‘Shield Immunity’ in Reducing COVID-19 Epidemic Spread | 10.1101/2020.04.01.20049767 | medRxiv | 10.5 (2.89) | 10.1038/s41591-020-0895-3 |
| 26 | Maximum entropy method for estimating the reproduction number: An investigation for COVID-19 in China | 10.1101/2020.03.14.20035659 | medRxiv | 7 (4.97) | 10.1103/physreve.102.032136 |
| 27 | Minimizing Population Health Loss in Times of Scarce Surgical Capacity | 10.1101/2020.07.26.20157040 | medRxiv | 13.25 (2.87) | 10.1016/j.jval.2022.01.027 |
| 28 | Modeling COVID-19: Forecasting and analyzing the dynamics of the outbreak in Hubei and Turkey | 10.1101/2020.04.11.20061952 | medRxiv | 8.75 (2.99) | 10.1002/mma.8181 |
| 29 | Negligible Risk of the COVID-19 Resurgence Caused by Work Resuming in China (outside Hubei): a Statistical Probability Study | 10.1101/2020.03.26.20044271 | medRxiv | 6.25 (3.1) | 10.1093/pubmed/fdaa046 |
| 30 | Parameter estimation and prediction for coronavirus disease outbreak 2019 (COVID-19) in Algeria | 10.3934/publichealth.2020026 | *AIMS Public Health* | 5 (2.45) |  |
| 31 | Predicting COVID-19 peaks around the world | 10.1101/2020.04.24.20078154 | medRxiv | 4.5 (2.52) | 10.3389/fphy.2020.00217 |
| 32 | Predicting the cumulative number of cases for the COVID-19 epidemic in China from early data | 10.1101/2020.03.11.20034314 | medRxiv | 5 (2.71) | 10.3934/mbe.2020172 |
| 33 | Prediction of COVID-19 spreading profiles in South Korea, Italy and Iran by data-driven coding | 10.1371/journal.pone.0234763 | *PLoS ONE* | 5.25 (1.5) |  |
| 34 | Progression of COVID-19 in Indian States - Forecasting Endpoints Using SIR and Logistic Growth Models | 10.1101/2020.05.15.20103028 | medRxiv | 8.25 (2.06) |  |
| 35 | Projections for first-wave COVID-19 deaths across the US using social-distancing measures derived from mobile phones | 10.1101/2020.04.16.20068163 | medRxiv | 11.25 (1.71) |  |
| 36 | Risk for COVID-19 infection and death among Latinos in the United States: examining heterogeneity in transmission dynamics | 10.1016/j.annepidem.2020.07.007 | *Annals of Epidemiology* | 10.75 (3.2) |  |
| 37 | Self-Burnout – A New Path to the End of COVID-19 | 10.1101/2020.04.17.20069443 | medRxiv | 3 (2.71) |  |
| 38 | Social distancing to slow the U.S. COVID-19 epidemic: interrupted time-series analysis | 10.1101/2020.04.03.20052373 | medRxiv | 7.25 (3.2) | 10.1371/journal.pmed.1003244 |
| 39 | Spatial analysis of COVID-19 spread in Iran: Insights into geographical and structural transmission determinants at a province level | 10.1101/2020.04.19.20071605 | medRxiv | 10 (2.45) | 10.1371/journal.pntd.0008875 |
| 40 | Spatial variability in the risk of death from COVID-19 in 20 regions of Italy | 10.1101/2020.04.01.20049668 | medRxiv | 9.5 (1.73) | 10.5588/ijtld.20.0262 |
| 41 | Study of Epidemiological Characteristics and In-silico Analysis of the Effect of Interventions in the SARS-CoV-2 Epidemic in India | 10.1101/2020.04.05.20053884 | medRxiv | 11 (6.06) |  |
| 42 | The contribution of pre-symptomatic infection to the transmission dynamics of COVID-2019 | 10.12688/wellcomeopenres.15788.1 | *Wellcome Open Research* | 11.5 (4.65) |  |
| 43 | The first three months of the COVID-19 epidemic: Epidemiological evidence for two separate strains of SARS-CoV-2 viruses spreading and implications for prevention strategies | 10.1101/2020.03.28.20036715 | medRxiv | 7.5 (4.65) | 10.7759/cureus.29146 |
| 44 | The spatio-temporal epidemic dynamics of COVID-19 outbreak in Africa | 10.1101/2020.04.21.20074435 | medRxiv | 6.75 (0.5) | 10.1017/S0950268820001983 |
| 45 | The time scale of asymptomatic transmission affects estimates of epidemic potential in the COVID-19 outbreak | 10.1101/2020.03.09.20033514 | medRxiv | 6.25 (0.5) | 10.1016/j.epidem.2020.100392 |
| 46 | Why lockdown? Simplified arithmetic tools for decision-makers, health professionals, journalists, and the general public to explore containment options for the novel coronavirus | 10.1101/2020.04.15.20066845 | medRxiv | 12 (0.58) | 10.1016/j.idm.2020.06.006 |

*Abbreviations:* DOI, digital object identifier; SD, standard deviation
