## Additional File 6 for "Inter-rater reliability of the Infectious Disease Modeling Reproducibility Checklist (IDMRC) as applied to COVID-19 computational modeling research"

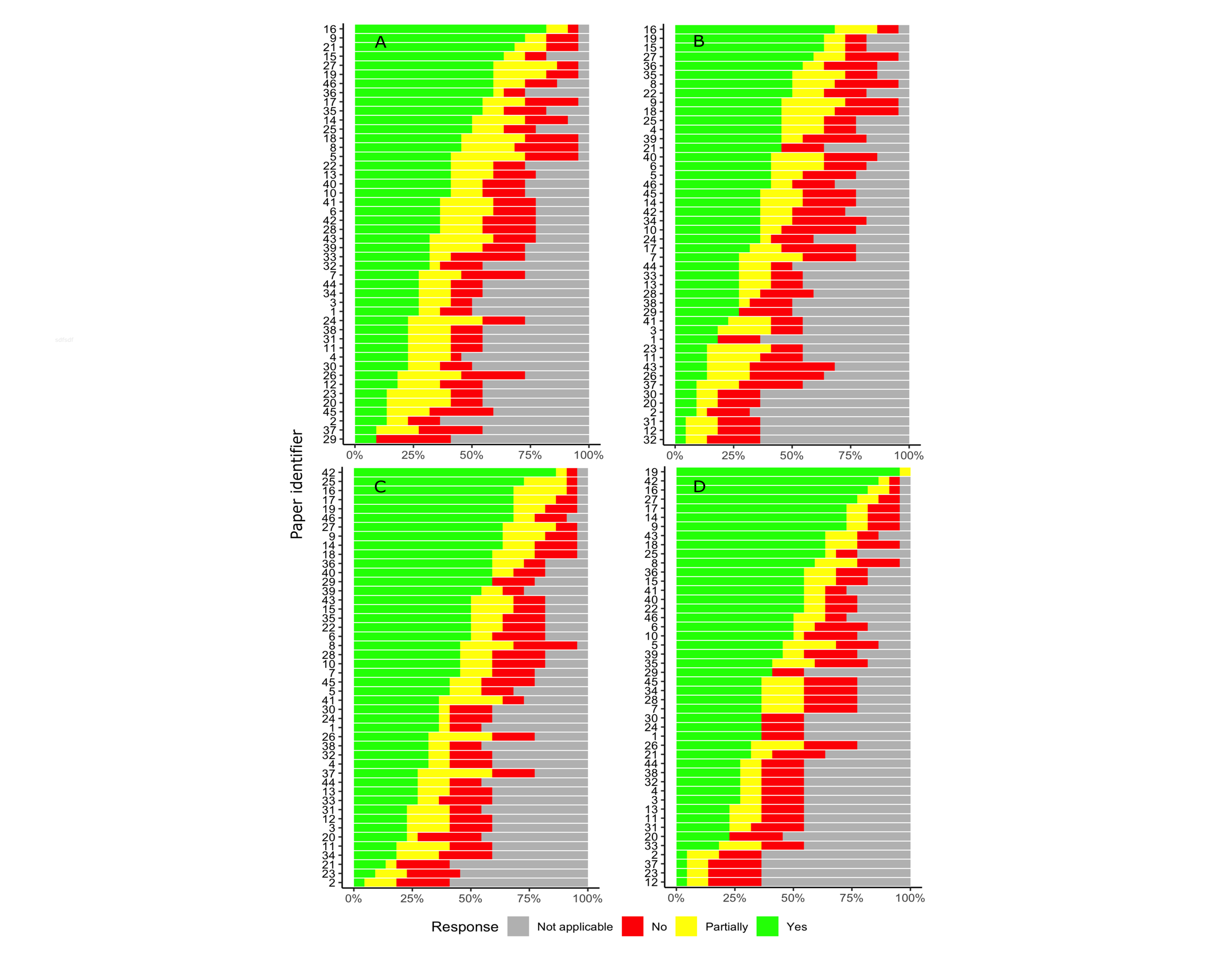

**Figure S1. Quantitative paper ranking (n = 46) among four reviewers.** (A-B) Rankings by new reviewers, KC and MM, respectively. (C-D) Rankings by reviewers with experience using the checklist, AAQ and DP, respectively. Green bars correspond to the number of reported elements in each publication (“yes” responses); yellow indicates partially reported elements; red indicates not reported elements, gray indicates not applicable responses.
