## Additional File 7 for "Inter-rater reliability of the Infectious Disease Modeling Reproducibility Checklist (IDMRC) as applied to COVID-19 computational modeling research"

**Table S6.** Publications (n = 46) ranked as being in the top (or bottom) 25% by all four reviewers are bolded.

|  | Reviewer | | | |
| --- | --- | --- | --- | --- |
| Publication Rank | KM | MM | AAQ | DP |
| Top 25% Ranked Publications | | | | |
| 1 | **16** | **16** | 42 | **19** |
| 2 | **9** | 15 | **25** | 42 |
| 3 | 21 | **19** | **16** | **16** |
| 4 | 15 | **27** | 17 | **27** |
| 5 | **27** | **36** | **19** | **9** |
| 6 | **19** | 35 | 46 | 14 |
| 7 | 46 | 8 | **27** | 17 |
| 8 | **36** | 22 | **9** | 43 |
| 9 | 17 | **9** | 14 | 18 |
| 10 | 35 | 18 | 18 | **25** |
| 11 | 14 | 4 | **36** | 8 |
| 12 | **25** | **25** | 40 | 15 |
| 13 | 18 | 39 | 29 | **36** |
| Bottom 25% Ranked Publications | | | | |
| 34 | 4 | 3 | **37** | 4 |
| 35 | **11** | 1 | 44 | 32 |
| 36 | **31** | **23** | 13 | 38 |
| 37 | 38 | **11** | 33 | 44 |
| 38 | 30 | 26 | **31** | **11** |
| 39 | 26 | 43 | 3 | 13 |
| 40 | **12** | **37** | **12** | **31** |
| 41 | **20** | **20** | **20** | **20** |
| 42 | **23** | 30 | **11** | 33 |
| 43 | 45 | **2** | 34 | **2** |
| 44 | **2** | **12** | 21 | **12** |
| 45 | **37** | **31** | **23** | **23** |
| 46 | 29 | 32 | **2** | **37** |
